# Multi-contrast magnetic resonance imaging of visual white matter pathways in glaucoma patients

**DOI:** 10.1101/2021.06.29.21259608

**Authors:** Shumpei Ogawa, Hiromasa Takemura, Hiroshi Horiguchi, Atsushi Miyazaki, Kenji Matsumoto, Yoichiro Masuda, Keiji Yoshikawa, Tadashi Nakano

**Affiliations:** Department of Ophthalmology, The Jikei University School of Medicine, Tokyo, Japan; Center for Information and Neural Networks (CiNet), Advanced ICT Research Institute, National Institute of Information and Communications Technology, Suita, Japan; Graduate School of Frontier Biosciences, Osaka University, Suita, Japan; Global Education Center, Waseda University, Tokyo, Japan; Brain Science Institute, Tamagawa University, Machida, Japan; Yoshikawa Eye Clinic, Machida, Japan

**Author notes:** **Corresponding authors**: Shumpei Ogawa, Department of Ophthalmology, Jikei University School of Medicine, 3-25-8, Nishi-Shimbashi, Minato-ku, Tokyo 105-8461 JAPAN, Hiromasa Takemura, Center for Information and Neural Networks (CiNet), Advanced ICT Research Institute, National Institute of Information and Communications Technology, 1-4 Yamadaoka, Suita-shi, Osaka 565-0871 JAPAN. These authors contributed equally (Shumpei Ogawa, Hiromasa Takemura). **Commercial relationship**: None. **Ethics statement**: This study was approved by the ethics committee of the Jikei University School of Medicine, National Institute of Information and Communications. Technology, and Tamagawa University. This study design followed the tenets of the Declaration of Helsinki. **Author contributions**: Designed the experiment: SO HT HH YM TN. Conducted the experiment: SO HH AM KM YM KY. Analyzed/interpreted data: SO HT HH. Wrote the article: SO HT HH. Proofed/revised article: SO HT HH AM KM YM KY TN.

**Keywords:** glaucoma, white matter, diffusion MRI, quantitative T1, optic radiation

## Abstract

**Purpose:** Glaucoma is a disorder that involves visual field loss caused by retinal ganglion cell damage. Previous diffusion magnetic resonance imaging (dMRI) studies have demonstrated that retinal ganglion cell damage affects tissues in the optic tract (OT) and optic radiation (OR). However, because previous studies have used a simple diffusion tensor model to analyze dMRI data, the microstructural interpretation of white matter tissue changes remains uncertain. In this study, we used a multi-contrast MRI approach to further clarify the type of microstructural damage that occurs in glaucoma patients.

**Methods:** We collected multi-shell dMRI data from 17 glaucoma patients and 30 controls using 3T MRI. Using the dMRI data, we estimated three types of tissue property metrics: intracellular volume fraction (ICVF), orientation dispersion index (ODI), and isotropic volume fraction (IsoV). Quantitative T1 (qT1) data, which may be relatively specific to myelin, were collected from all subjects.

**Results:** In the OT, all four metrics showed significant differences between the glaucoma and control groups. In the OR, only the ICVF showed significant between-group differences. ICVF was significantly correlated with qT1 in the OR of glaucoma patients, although qT1 did not show any abnormality at the group level.

**Conclusions:** Our results suggest that at the group level, tissue changes in the glaucoma patients’ OR might be explained by axonal damage, which is reflected in the intracellular diffusion signals, rather than myelin damage. The significant correlation between ICVF and qT1 suggests that myelin damage might also occur in a smaller number of severe cases.

## Precis

While previous studies have revealed that glaucoma affects white matter, a precise understanding of microstructural abnormalities remains to be achieved. This work demonstrates that glaucoma affects intracellular diffusion properties along the optic radiation using multi-contrast MRI.

Glaucoma is a chronic progressive optic neuropathy in which retinal ganglion cell damage causes visual field loss and optic nerve damage (Figure 1A) ^1^. Because glaucoma is the leading cause of blindness with a high prevalence rate in elderly populations ^2^, there is an urgent need to understand how glaucoma affects nerve fiber pathways and brain areas ^3^.

**Figure 1.**
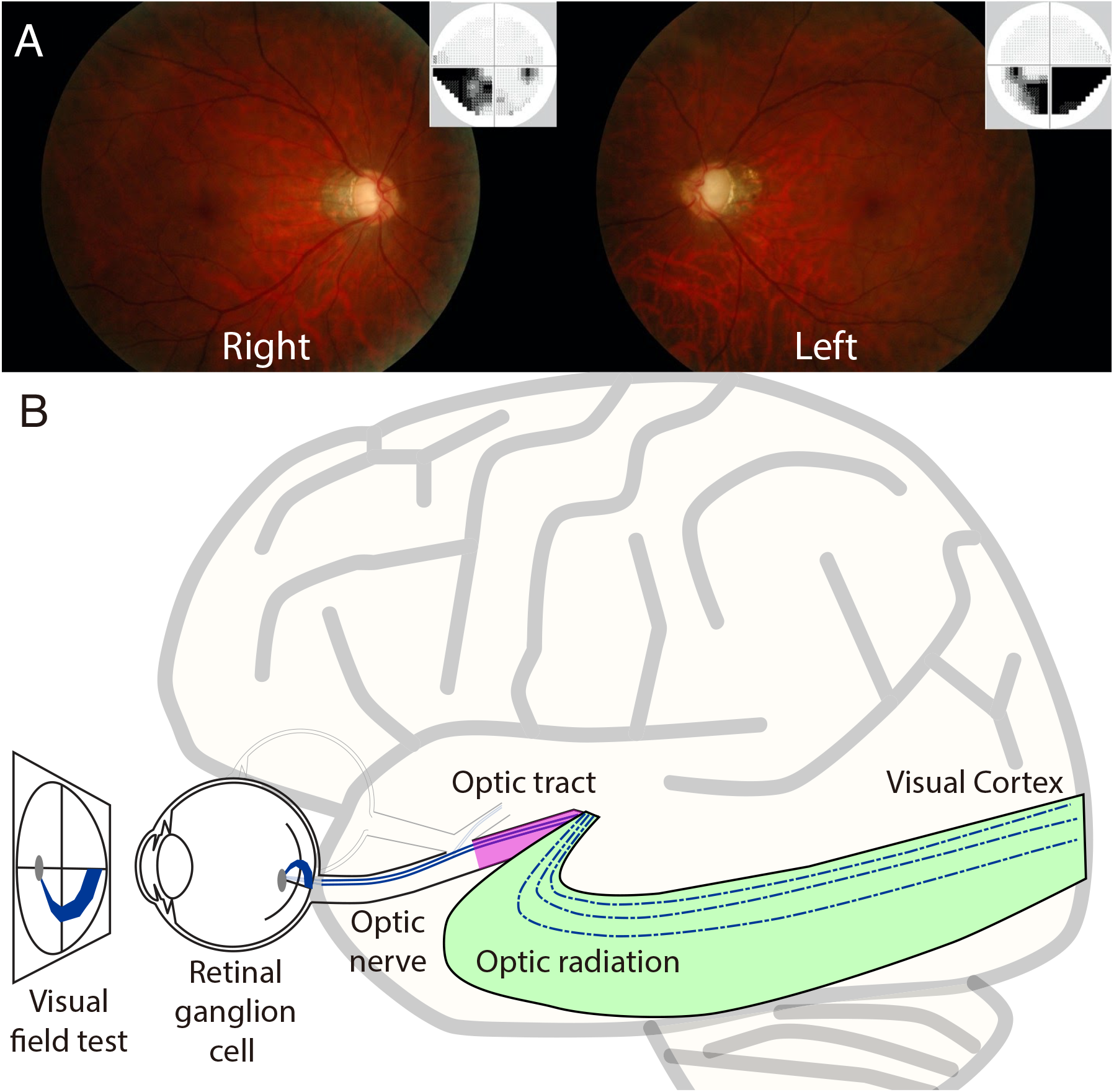
Clinical features of glaucoma patients and schematic illustration of the visual system. **A**. Glaucomatous fundus photograph of a representative glaucoma patient (Glc-015; *left panel*, right eye; *right panel*, left eye). The small panel at the top right of each main panel depicts associated visual field loss, as measured using the Humphrey field analyzer (Carl Zeiss Meditec, Dublin, CA). The superior neural losses correspond to deficits on the lower visual fields of both eyes. **B**. A schematic illustration of the early visual pathways from the eye to the primary visual cortex. Retinal ganglion cells receive visual information from photoreceptor cells via two types of intermediate cells. The optic nerve is composed of retinal ganglion cell axons and glial cells, which extend from the optic disc to the optic chiasm and continue as the optic tract to the lateral geniculate nucleus (LGN). From the LGN, fibers of the optic radiation carry visual information to the primary visual cortex in the occipital lobe of the brain.

Recently, diffusion tensor imaging (DTI) has enabled researchers to investigate tissue abnormalities of visual white matter tracts, such as the optic tract (OT) and optic radiation (OR; Figure 1B), in living humans ^4,5^. DTI studies have demonstrated that glaucoma patients show abnormalities in diffusivity measurements, such as fractional anisotropy (FA) ^6^, along these tracts ^7–18^. However, a major limitation is that DTI-based metrics do not directly correlate with specific types of tissue properties, such as axon diameter and myelination ^5,19–23^.

This study aimed to improve our understanding of the white matter consequences of glaucoma by combining two different neuroimaging approaches. One approach is neurite density and orientation dispersion imaging (NODDI ^24^), which uses a biophysical model to provide several distinct metrics (intracellular volume fraction [ICVF], orientation dispersion index [ODI], and isotropic volume fraction [IsoV]) from diffusion magnetic resonance imaging (dMRI) data. For example, ICVF aims to quantify the volume fraction of cylinders designed to describe axons and dendrites based on restricted diffusion signals. NODDI-based metrics have been hypothesized to be more specific to the properties of axons, than DTI-based metrics ^25–28^. The other approach uses qT1, which quantifies the T1 relaxation time by combining structural MRI data acquired with multiple parameters ^29–31^. qT1 is hypothesized to be relatively specific to myelin levels in the white matter based on comparisons with histological data ^29–31^. We investigated white matter tissue properties in glaucoma patients and age-matched controls using NODDI and qT1, since they are hypothesized to be correlated with different types of microstructural properties and can be measured by MRI sequences available for clinical studies and publicly available software.

## Materials and Methods

This study was approved by the ethics committee of the Jikei University School of Medicine, National Institute of Information and Communications Technology, and Tamagawa University. All subjects provided written informed consent to participate in the study. The study design followed the tenets of the Declaration of Helsinki. The data and code for reproducing figures and statistical analyses will become available at [URL will be inserted upon acceptance of the manuscript].

### Subjects

A total of 17 glaucoma patients (Table 1; eight women; mean age, 56.6 years; age range, 24–72 years) participated in this study. Thirty volunteers with normal visual function and no visual field defects also participated in this study as control subjects (Table 2; 14 women; mean age, 51.4 years; age range, 36–71 years). With reference to a previous study ^32^, this sample size was predicted to be sufficient to identify a large effect size in group difference (d’ = 1.10) using a two-tailed two-sample t-test (evaluated by G*Power version 3.1.9.6).

**Table 1.** Glaucoma patients’ profile. HFA: Humphrey visual field analyzer, POAG: primary open angle glaucoma, NTG: normal tension glaucoma, PEXG: pseudoexfoliation glaucoma, SOAG: secondary open angle glaucoma, MD: mean deviation.

|  | Sex | R-HFA(MD) | L-HFA(MD) | Disease type | Age |
| --- | --- | --- | --- | --- | --- |
| Glc-001 | F | -27.95 | -4.62 | POAG | 70-75 |
| Glc-002 | F | -9.48 | -14.27 | NTG | 56-60 |
| Glc-003 | M | -13.22 | -12.95 | NTG | 51-55 |
| Glc-004 | F | -25.92 | -31.14 | PEXG | 61-65 |
| Glc-005 | F | -11.36 | -14.02 | NTG | 41-45 |
| Glc-006 | F | -13.35 | -14.02 | NTG | 61-65 |
| Glc-007 | F | -15.55 | -24.13 | SOAG | 70-75 |
| Glc-008 | M | 1.76 | 0.38 | NTG | 56-60 |
| Glc-009 | M | -2.55 | -14.96 | POAG | 66-70 |
| Glc-010 | M | 1.08 | -2 | NTG | 61-65 |
| Glc-011 | F | -10.09 | 0.46 | NTG | 51-55 |
| Glc-012 | M | -11.65 | -0.52 | NTG | 51-55 |
| Glc-013 | M | -8.42 | -14.37 | POAG | 46-50 |
| Glc-014 | M | -2.18 | -8.67 | NTG | 21-25 |
| Glc-015 | M | -13.42 | -17.39 | POAG | 41-45 |
| Glc-016 | F | -12.94 | -20.25 | NTG | 51-55 |
| Glc-017 | M | -10.06 | -21.81 | NTG | 56-60 |

**Table 2.** Control subjects’ profile.

|  | Age | Sex |
| --- | --- | --- |
| Ctl-001 | 36-40 | F |
| Ctl-002 | 41-45 | F |
| Ctl-003 | 36-40 | F |
| Ctl-004 | 36-40 | M |
| Ctl-005 | 70-75 | M |
| Ctl-006 | 66-70 | F |
| Ctl-007 | 66-70 | F |
| Ctl-008 | 41-45 | F |
| Ctl-009 | 46-50 | M |
| Ctl-010 | 36-40 | M |
| Ctl-011 | 46-50 | M |
| Ctl-012 | 51-55 | M |
| Ctl-013 | 41-45 | M |
| Ctl-014 | 56-60 | M |
| Ctl-015 | 41-45 | M |
| Ctl-016 | 56-60 | F |
| Ctl-017 | 51-55 | F |
| Ctl-018 | 51-55 | F |
| Ctl-019 | 46-50 | F |
| Ctl-020 | 51-55 | F |
| Ctl-021 | 56-60 | F |
| Ctl-022 | 51-55 | M |
| Ctl-023 | 61-65 | F |
| Ctl-024 | 36-40 | M |
| Ctl-025 | 51-55 | F |
| Ctl-026 | 51-55 | M |
| Ctl-027 | 66-70 | M |
| Ctl-028 | 41-45 | M |
| Ctl-029 | 56-60 | M |
| Ctl-030 | 56-60 | M |

### Clinical features of the patients with glaucoma

Patients with glaucoma who participated in this study were diagnosed with open-angle glaucoma (primary and secondary open-angle glaucoma) by experienced ophthalmologists at the Department of Ophthalmology, Jikei University School of Medicine. All patients underwent a comprehensive ophthalmologic examination, including measurement of visual fields using the Humphrey Visual Field Analyzer (HFA) 24-2 or 30-2 Swedish interactive thresholding algorithm standard program by HFA (Carl Zeiss Meditec, Dublin, CA), and measurement of retinal nerve fiber layer thickness using Cirrus HD-optical coherence tomography (Carl Zeiss Meditec, Dublin, CA) or Spectralis optical coherence tomography (Version 4; Heidelberg Engineering, Heidelberg, Germany). The diagnosis of glaucoma was based on glaucomatous optic neuropathy and visual field defects consistent with optic changes ^33^. Figure 1A depicts the fundus photograph and visual field loss in a representative glaucoma patient. Results of HFA in all patients were described in Table 1, in a unit of mean deviation from healthy populations.

### MRI data acquisition

MRI images were acquired using a Siemens 3T MAGNETOM Trio Tim scanner with a 32 channel head coil at the Tamagawa University Brain Science Institute, Machida, Japan.

#### Structural MRI data acquisition

T1-weighted MP-RAGE images were collected from all subjects (1 mm isotropic voxels; repetition time [TR], 2000 ms; echo time [TE], 1.98 ms). Acquisition of the T1-weighted MRI data took 9 min and 18 s per subject.

T2-weighted SPACE images were also acquired from 42 subjects (all 17 glaucoma patients and 25 control subjects; 1 mm isotropic voxels; TR, 3000 ms; TE, 478 ms) to obtain T1-weighted/T2-weighted ratio maps (T1w/T2w) ^34^. Acquisition of T2-weighted MRI data took approximately 11 min and 14 s per subject.

#### dMRI data acquisition

dMRI data were collected from all subjects using single-shot spin-echo, echo planar imaging (EPI; 32 directions with b = 700 s/mm^2^; 64 directions with b = 2000 s/mm^2^; 1.7 mm isotropic voxels; TR, 4500 ms; TE, 94 ms; in-plane acceleration, 2; multi-band factor, 3; phase partial Fourier, 6/8; diffusion scheme, monopolar) implemented in a multi-band accelerated EPI pulse sequence provided by the Center for Magnetic Resonance Research, Department of Radiology, University of Minnesota; https://www.cmrr.umn.edu/multiband/) ^35^. Twelve non-diffusion-weighted (*b* = 0) measurements were also acquired. To minimize the impact of EPI distortion in subsequent analyses, two image sets were acquired with reversed phase-encoding directions (A-P and P-A). Acquisition of the dMRI data took approximately 9 min and 11 s per subject.

#### qT1 data acquisition

For all subjects, qT1 was measured following protocols described in previous publications ^29,32^. Four fast low-angle shot (FLASH) images with flip angles of 4°, 10°, 20°, and 30° (TR, 12 ms; TE, 2.41 ms) and isotropic 2-mm voxels were acquired. Five additional spin-echo inversion-recovery (SEIR) scans with an EPI readout (TR, 3000 ms; TE, 49 ms; 2× acceleration) were acquired to remove field inhomogeneities. The inversion times were 50, 200, 400, 1200, and 2400 ms. The in-plane resolution and slice thickness of the additional scans were 2 × 2 mm and 4 mm, respectively. Acquisition of qT1 data took approximately 13 min and 30 s per subject.

### MRI data analysis

#### Structural MRI data preprocessing

T1-weighted images of individual subjects were aligned to the anterior commissure– posterior (AC-PC) space. This aligned image on the AC-PC coordinate was then used as a common coordinate frame across the dMRI and qT1 datasets.

T1w/T2w maps were also calculated by aligning the T2-weighted image with the T1-weighted image in the AC-PC space and calculating the ratio between the image intensities of the T1-weighted image and T2-weighted image for all 42 subjects who participated in the T2-weighted image acquisition.

#### dMRI data preprocessing

dMRI data were preprocessed using the TOPUP and EDDY tools in FSL (https://fsl.fmrib.ox.ac.uk/fsl/fslwiki) to correct for susceptibility-induced distortion and eddy-current artifacts ^36,37^. The dMRI data were then aligned to the T1-weighted images in the AC-PC space using a 14-parameter constrained nonlinear coregistration algorithm. The tensor model was then fitted to the dMRI data using a least-squares algorithm to estimate FA, mean diffusivity (MD), axial diffusivity (AD), and radial diffusivity (RD). Furthermore, NODDI was also fitted to the dMRI data to obtain ICVF, ODI, and IsoV (Figure 2B), using the NODDI MATLAB toolbox (http://mig.cs.ucl.ac.uk/index.php?n=Tutorial.NODDImatlab).

**Figure 2.**
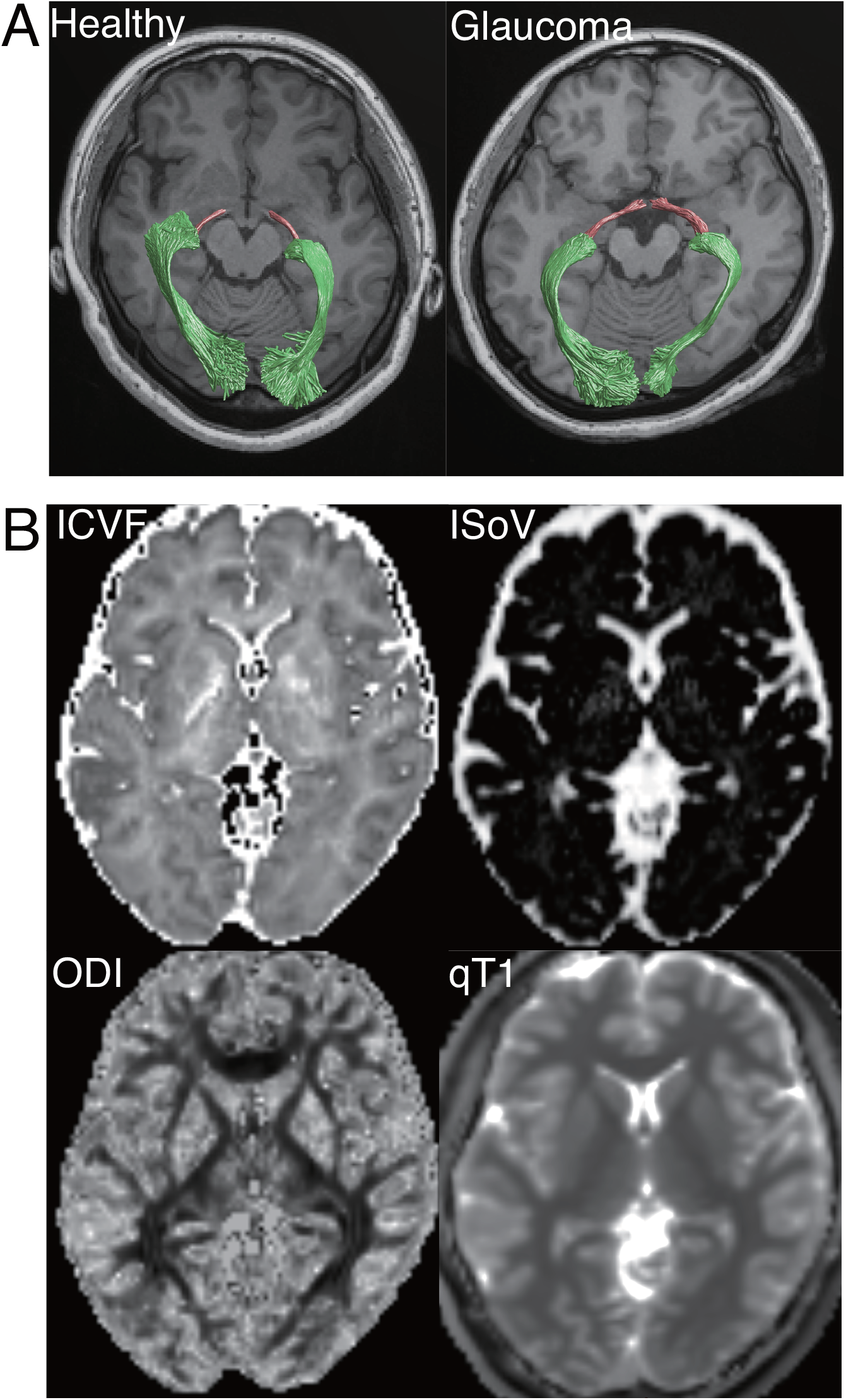
**A**. Visual white matter tracts identified by probabilistic tractography in representative subjects in each group (left, Ctl-008; right, Glc-004; magenta, OT; green, OR), overlaid on an axial slice of a T1-weighted image, located inferior to the tracts. **B**. Tissue measurement maps (ICVF, ODI, IsoV, and qT1) in a representative healthy subject (Ctl-008).

#### qT1 data preprocessing

Both the FLASH and SEIR scans were processed using the mrQ software package (https://github.com/mezera/mrQ) to produce qT1 maps (Figure 2B) ^29^, and the qT1 maps were registered to the T1-weighted images using the FSL FLIRT tool ^38^.

#### Tract identification and evaluation

*Tract identification*. We identified the OT and OR by analyzing the dMRI data using the same method as that used in previous studies (Figure 2A) ^32,39^. Briefly, tracking was performed between regions of interest, which were defined manually (lateral geniculate nucleus [LGN]) ^32^ or by FreeSurfer segmentation (optic chiasm and primary visual cortex [V1]) ^40–42^. Outlier streamlines were excluded using Automated Fiber Quantification (AFQ; https://github.com/yeatmanlab/AFQ) ^43^.

##### Evaluation tissue properties

We evaluated the tissue properties of OT and OR using AFQ. Briefly, each streamline was resampled to 100 equidistant nodes, and tissue properties (qT1, ICVF, ODI, and IsoV) were calculated at each node of each streamline. The properties at each node were summarized by taking a weighted average of the tissue measurements (qT1, ICVF, ODI, and IsoV) on each streamline within that node. The weight of each streamline was based on the Mahalanobis distance from the tract core. Data from the left and right hemispheres were averaged. We excluded the first and last 10 nodes because they are susceptible to crossing with U-fibers and partial voluming with gray matter. While the results of the remaining 80 nodes are plotted as tract profiles (Figures 3-4), we averaged the data of 80 nodes to obtain a single-number summary of each metric per subject for statistical comparisons. For the dMRI metrics, we report the average results of the two runs.

**Figure 3.**
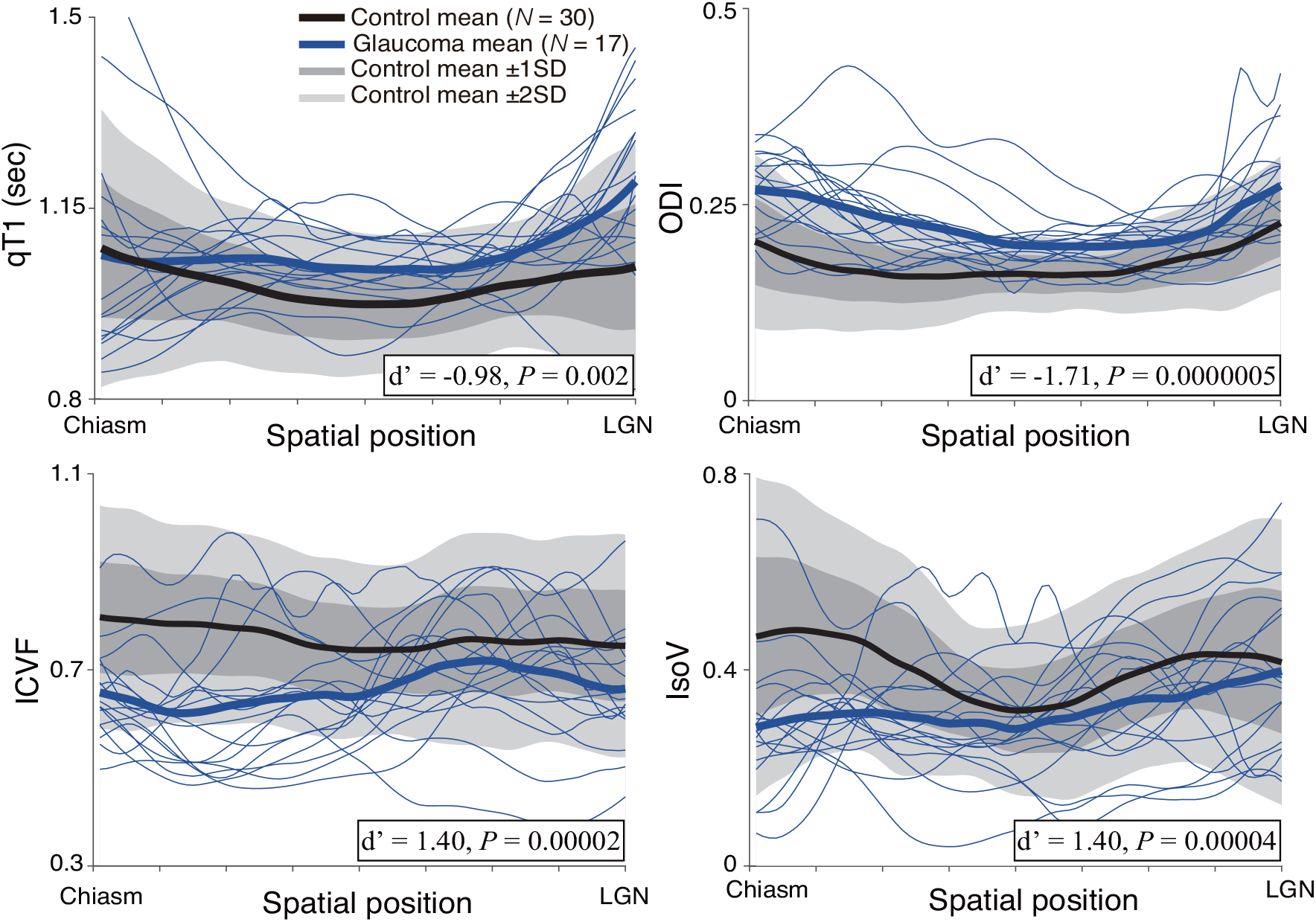
Tissue properties along the OT. The four panels show the qT1 (left upper panel), ICVF (left lower panel), ODI (right upper panel), and IsoV (right lower panel) measurements along the OT. Profiles of individual glaucoma patients are depicted as thin blue curves. Thick curves show the mean of each group (glaucoma: blue, healthy control: black). The lighter gray shades indicate a range of ± 2 S.D. from the control mean, and the darker gray band shows ± 1 S.D. from the control mean. The horizontal axis describes the normalized position along the tract (left: anterior, right: posterior).

##### Tissue properties controlled for age

Linear regression was performed to predict the tissue properties (qT1, ICVF, ODI, and IsoV) along the OT and OR according to the ages of the subjects, which were pooled across both glaucoma and control groups. We then calculated the residual, which is the difference between the measured tissue properties and tissue properties predicted by age ^44^. We used the residual as the tissue property controlled for age to evaluate the impact of age variability on the main analysis.

#### Statistical analyses

##### Group comparison

Inter-group differences in the major metrics of interest (qT1, ICVF, ODI, and IsoV) were assessed using a two-tailed two-sample t-test. We defined the significance level (α) as 0.05. The effect sizes (Cohen’s d’) of the group differences were calculated for each comparison, including those of other metrics (FA, MD, MTV, and T1w/T2w).

##### Correlation between measurements in glaucoma patients

We also evaluated between-metric correlations in the extent of the glaucoma patient’s tissue abnormality in each metric (qT1, ICVF, ODI, and IsoV) in the OT and OR. To this end, we first calculated the degree of deviation from the control mean in each glaucoma patient in units of standard deviation of the control subjects. We then calculated the inter-patient Pearson correlation of each metric in the same tract and evaluated its statistical significance. α was defined as 0.008, which is equivalent to *P* = 0.05, with Bonferroni correction for six comparisons.

##### Correlation between MRI measurements and visual field tests

We also assessed the correlation between tissue properties of the OT and the OR and the visual field test scores (mean deviation value of HFA) in patients with glaucoma. To this end, we normalized the MRI measurements of the glaucoma patients by calculating the degree of deviation from the control mean, using units of standard deviation in the control subjects. The visual field test scores of each glaucoma patient (Table 1) were averaged across the two eyes. We then calculated the Pearson correlation between the MRI measurements and the visual field test results in glaucoma patients. For OR, we only focused on the ICVF since it is the only metric showing significant abnormality in glaucoma patients (Figure 4).

**Figure 4.**
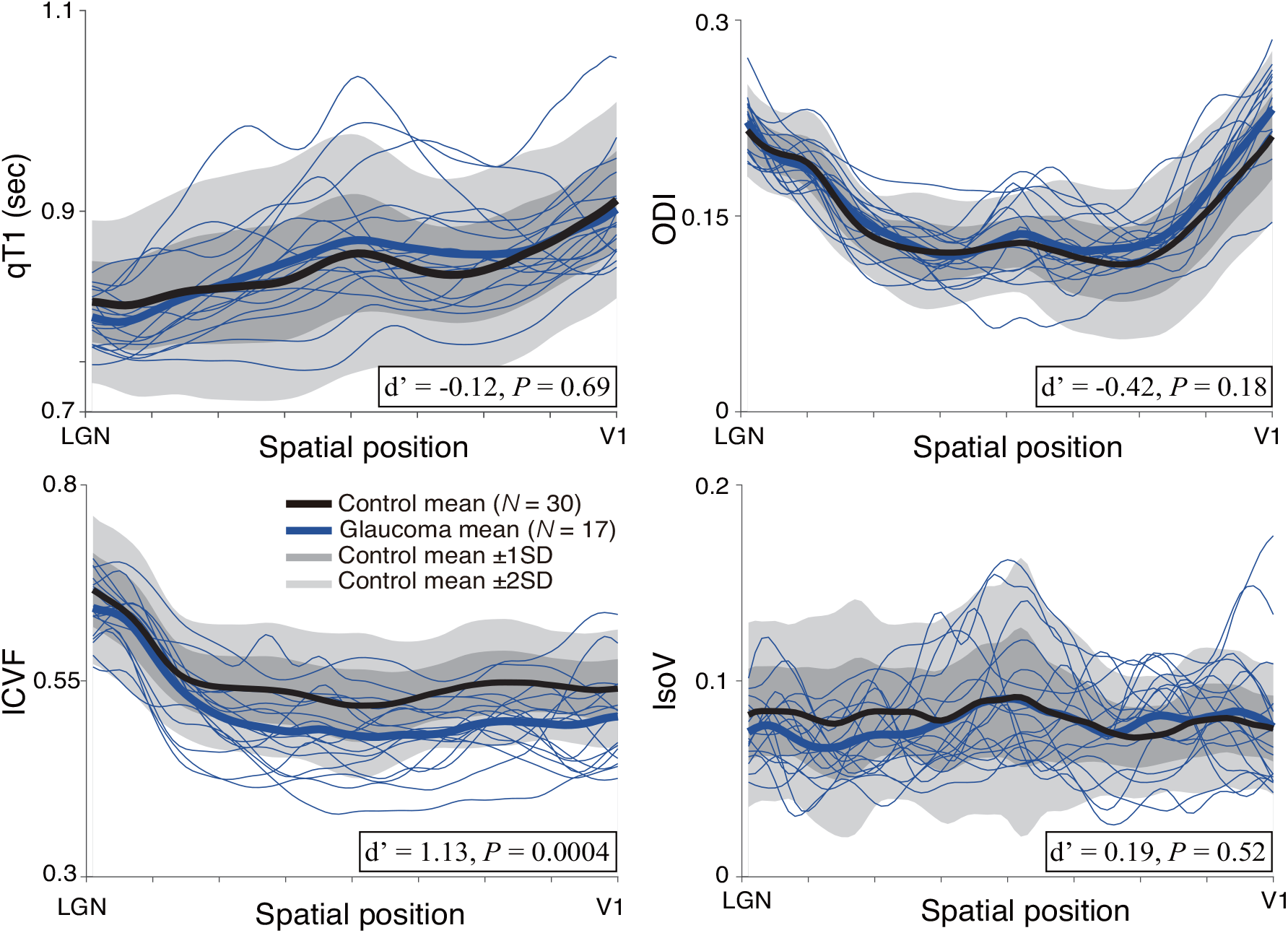
Tissue properties along the OR (left upper panel, qT1; left lower panel, ICVF; right upper panel, ODI; right lower panel, IsoV). Other conventions are identical to those of Figure 3.

## Results

We successfully identified the OT and OR in all hemispheres (see Figure 2A for representative subjects). We then calculated the four tissue measurements (ICVF, ODI, IsoV, and qT1) along the OT and OR.

### Glaucoma affected all types of tissue measurements in the OT

Figure 3 depicts the profile of the OT in the control (black) and glaucoma (blue) subjects. The gray shaded regions in Figure 3 show the standard deviation from the control mean for qT1 (left upper panel), ICVF (left lower panel), ODI (right upper panel), and IsoV (right lower panel), along the length of the OT. The individual blue curves depict data from individual glaucoma patients. We found significant group differences in all measurements; the glaucoma patients showed significantly higher qT1 (two-tailed two-sample t-test; qT1, d’ = −0.98, t_45_ = −3.23, 95% confidence interval [CI], −0.09 to −0.02, *P* = 0.002), significantly lower ICVF (d’ = 1.40, t_45_ = 4.82, 95% CI, 0.06 to 0.15, *P* = 0.00002), significantly higher ODI (d’ = −1.71, t_45_ = −5.82, 95% CI, −0.07 to −0.03, *P* = 0.0000005), and significantly lower IsoV (d’ = 1.40, t_45_ = 4.55, 95% CI, 0.05 to 0.12, *P* = 0.00004). These differences were preserved after controlling for age (Supplementary Figure 1).

### Glaucoma affected ICVF in the OR, but not other measurements

Figure 4 shows the profile of the OR in the control and glaucoma subjects. We observed significantly lower ICVF in glaucoma patients (Figure 4; d’ = 1.13, t_45_ = 3.80, 95% CI, 0.02 to 0.06, *P* = 0.0004). By contrast, we did not find significant differences in qT1 (d’ = −0.12, t_45_ = −0.40, 95% CI, −0.03 to 0.02, *P* = 0.69), ODI (d’ = −0.42, t_45_ = −1.38, 95% CI, − 0.02 to 0.00, *P* = 0.18), or IsoV (d’ = 0.19, t_45_ = 0.66, 95% CI, −0.01 to 0.01, *P* = 0.52). Similar results were obtained when we controlled the tissue properties for age (Supplementary Figure 2).

### Do multiple MRI measurements detect similar types of tissue abnormalities within the same tract?

For each MRI metric along the OT, we quantified the deviation from the control mean for each glaucoma patient and calculated the correlation between them. We found a significant correlation between the extent of abnormalities in ICVF and IsoV (Figure 5A; R = 0.70; P = 0.002). The correlations between other pairs did not reach statistical significance (α= 0.008; qT1-ICVF, R = −0.45, P = 0.07; qT1-ODI, R = 0.30, P = 0.25; qT1-IsoV, R = −0.49; P = 0.05; ICVF-ODI, R = 0.02, P = 0.93; ODI-IsoV, R = −0.57, P = 0.02), suggesting that these metrics may reflect multiple underlying sources of tissue abnormalities in the OT.

**Figure 5.**
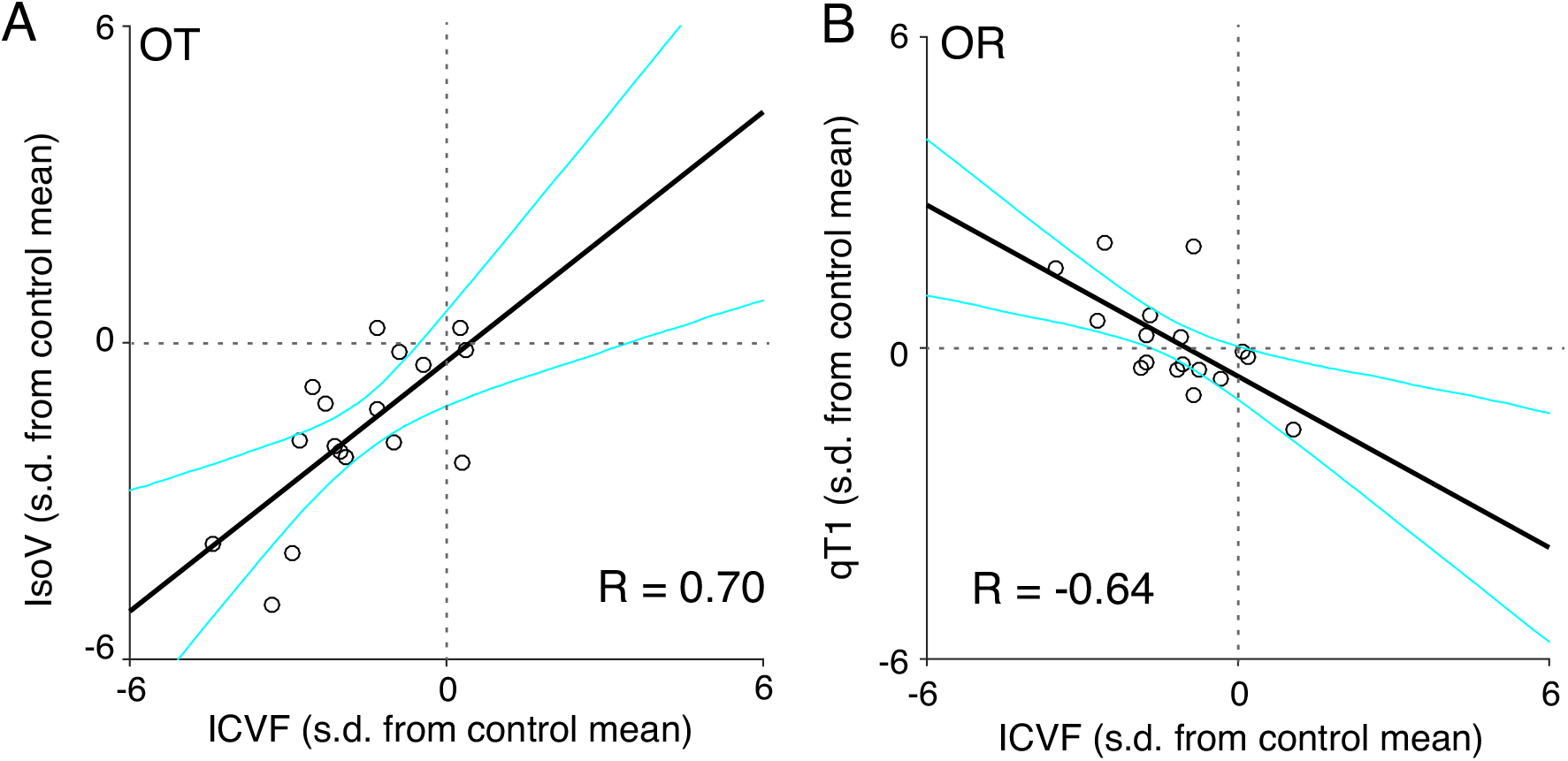
Correlations between multiple MRI measurements in glaucoma patients. The two-dimensional scatter plots depict the extent of deviation of each individual glaucoma patient (N = 17) from the control mean, with units of 1 S.D. of the control group. Individual dots are data points from individual glaucoma patients. Black thick lines depict regression lines, while cyan curves indicate the 95% confidence intervals of linear regression estimated by the bootstrapping method. **A**. Correlation between ICVF and IsoV in the OT. **B**. Correlation between ICVF and qT1 in the OR.

We also investigated the correlation between the extent of abnormalities in each MRI metric along the OR in glaucoma patients. We found a significant correlation between qT1 and ICVF (Figure 5B; R = −0.64; P = 0.005), while other pairings were not significantly correlated (qT1-ODI, qT1-IsoV, ICVF-ODI, ICVF-IsoV, and ODI-IsoV; R = − 0.33, 0.46, 0.39, −0.11, and 0.00; P = 0.19, 0.06, 0.13, 0.69 and 1.00, respectively).

### Relationship between white matter tissue measurement and visual field loss

We assessed the extent to which this inter-patient variability in MRI measurements could be explained by the severity of visual field loss by evaluating the correlations between MRI measurements and visual field test data (Table 1) among glaucoma patients. In the OT, none of the four MRI metrics showed a significant correlation with the visual field test (R = 0.33, −0.34, 0.32, and −0.31; P = 0.19, 0.18, 0.21, and 0.22; for qT1, ICVF, ODI, and IsoV, respectively). By contrast, we found that ICVF along the OR was significantly correlated with the visual field test (R = 0.50, P = 0.04; Supplementary Figure 3A), although this correlation was only marginally significant when controlling for differences in age (Supplementary Figure 3B; R = 0.43, P = 0.09). Therefore, the robustness of this correlation should be re-examined in future studies.

### Comparisons across multiple MRI-based tissue measurement metrics

Previous studies assessed the tissue properties of white matter tracts using DTI-based metrics (FA, MD, RD, and AD) ^6^. Furthermore, macromolecular tissue volume (MTV) ^29^, which has different sensitivities for lipids compared with qT1 ^45^, has also been used to evaluate white matter tissue properties ^46^. We evaluated the extent to which these metrics deviated between glaucoma patients and controls, to facilitate comparison of the present data with those of previous studies.

Figure 6 shows how each MRI metric in individual glaucoma patients deviated from the control mean. In this plot, the vertical axis indicates the effect size of the difference (d’) between individual glaucoma patients and the control mean for each metric. We found that FA was lower in glaucoma patients in both the OT and OR (effect size of group difference, d’ = 2.45 and 1.50 in OT and OR, respectively). We also found higher MD in glaucoma patients, although the difference in the OT was small and not consistent across all patients (d’ = −0.52 and −1.03 in OT and OR, respectively). In both OT and OR, we found much higher RD in the glaucoma patients (d’ = −1.63 and −1.44, respectively), although AD did not show a consistent pattern across them (d’ = 0.76 and −0.23, respectively). Therefore, profound differences in FA can be mostly explained by higher RD, rather than lower AD.

**Figure 6.**
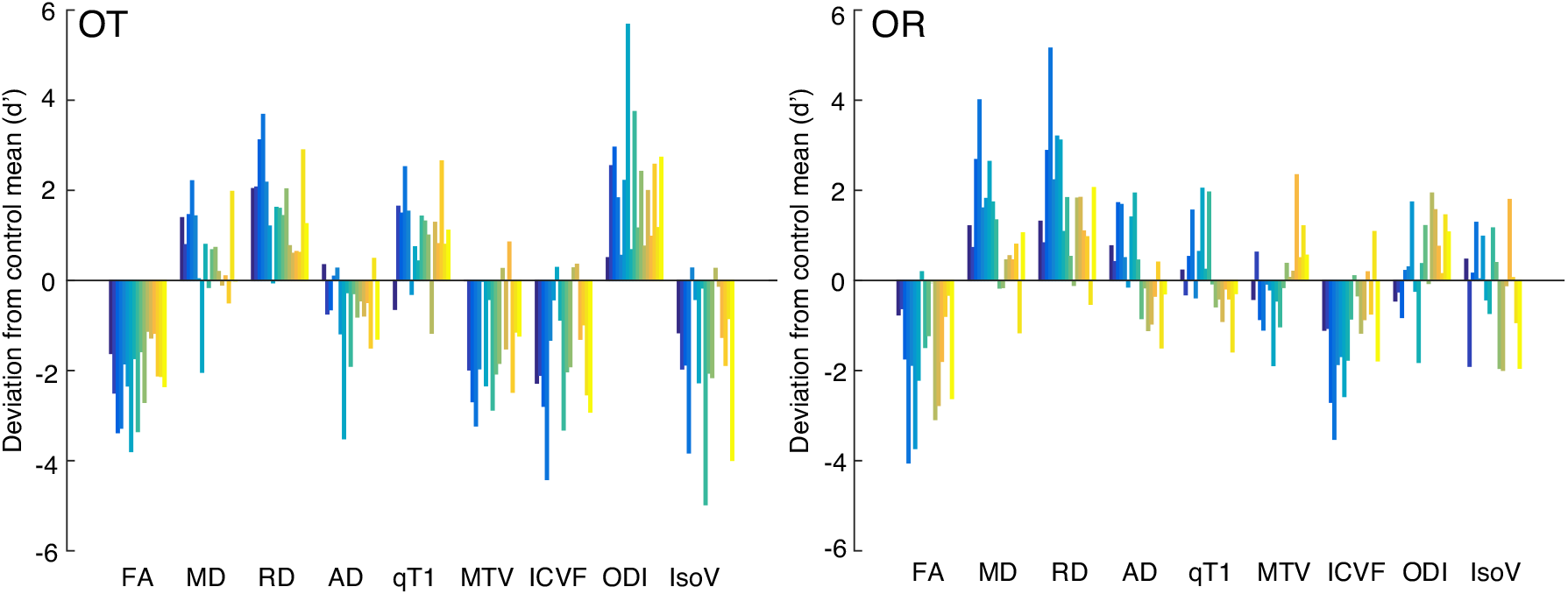
Evaluation of different MRI metrics derived from the diffusion tensor model, qMRI and NODDI. *Left panel*: OT. *Right panel*: OR. The vertical axis of each plot represents the extent to which individual glaucoma patients (color bars; Glc001-017) deviate from the control mean (*N* = 30). The vertical axis unit is the effect size (d’) of the difference between individual glaucoma patients (each colored bar) and the control mean.

In the OT, the glaucoma patients consistently showed lower MTV (d’ = 1.31). By contrast, we did not find consistently lower MTV among glaucoma patients (d’ = 0.02), similar to the observations on qT1 (d’ = −0.12). Therefore, similarly to our previous study on Leber’s hereditary optic neuropathy (LHON) ^32^, qT1 and MTV showed abnormalities in the OT and a lack of abnormalities in the OR.

We also evaluated the T1w/T2w ratio, which has been used in recent neuroimaging studies (Supplementary Figure 4) ^34,47,48^. In the OT, glaucoma patients showed slightly lower T1w/T2w than the controls (effect size of group difference, d’ = 0.72); however, in the OR, a group-level difference was almost absent (d’ = 0.04). In both tracts, two glaucoma patients showed higher T1w/T2w than the controls (Supplementary Figure 5; Glc-013 and 014). However, because these two patients did not show clear opposite trends in dMRI and quantitative MRI metrics (Figure 6), it is unclear whether these trends reflect a true tissue difference or measurement bias.

## Discussion

Previous anatomical studies on glaucoma patients and non-human primate models revealed that glaucoma causes tissue changes in the LGN ^49–52^ and V1 ^49^, indicating that glaucoma affects tissues of the subsequent visual areas receiving inputs from retinal ganglion cells. More recently, DTI studies have demonstrated abnormalities in diffusivity measurements (such as FA) along the OT and OR in glaucoma patients ^7,8,12,13,53–66^. However, the types of microstructural changes that occur as a consequence of glaucoma are not yet understood since DTI-based metrics are not specific to myelin damage or axonal loss ^20–22^.

We combined NODDI and qT1 to overcome the limitations of DTI. We found that all four MRI metrics (qT1, ICVF, ODI, and IsoV; Figure 3) indicated abnormalities in the OT of glaucoma patients. This suggests that OT tissue abnormalities in glaucoma may be caused by multiple factors. By contrast, only ICVF indicated abnormalities in the OR of glaucoma patients (Figure 4), which suggests that tissue abnormalities in the OR are associated with more specific neurobiological mechanisms. Because it is natural to interpret ICVF in white matter voxels as a measurement of apparent axonal density ^67^, our finding may indicate that glaucoma causes axonal damage in the OR, rather than myelin damage. However, this interpretation remains speculative because histological validation of ICVF along the OR remains to be performed.

We also found that ICVF was correlated with qT1 in the OR of glaucoma patients (Figure 5B), while on average, qT1 did not show any abnormalities (Figure 4). This result indicates that glaucoma patients with severe ICVF abnormalities tend to exhibit some degree of abnormality in qT1. While speculative, these results suggest the possibility that in the OR of glaucoma patients, axonal damage (quantified by ICVF) occurs earlier than myelin damage (quantified by qT1). Myelin damage may occur at a later stage of disease progression. This hypothesis should be tested in future longitudinal studies or preclinical MRI studies on animal models ^51^, in which control of the duration since disease onset is practical.

The DTI approach has been widely adapted to clinical groups other than glaucoma, such as amblyopia ^68–70^, LHON ^32,71^, retinitis pigmentosa ^72^, and macular degeneration ^71,73–75^. However, comparisons of each of these studies with the present results are not straightforward because all of these studies used a simplistic DTI-based approach. Among these previous studies, Takemura et al. (2019) used a similar combination of MRI methods (DTI and qT1) to evaluate white matter tissue properties in patients with LHON. Although their study did not use NODDI, we can compare our DTI results obtained from glaucoma patients (Figure 6) with those obtained from LHON patients. This comparison revealed consistent findings: 1) both diffusivity and qT1 demonstrated abnormalities in the patients’ OT, and 2) abnormal diffusivity measurements were made from the patient’s OR; however, qT1 measurements showed no evidence of abnormality, suggesting that a main finding may be generalizable to LHON and glaucoma, both of which cause retinal ganglion cell damage.

However, the data from our glaucoma patients showed a notable difference from LHON ^32^. Specifically, while the LHON study did not show a large abnormality in RD along the OT, the glaucoma data acquired in this study did (Figure 6), suggesting that glaucoma and LHON may cause different types of tissue changes. We speculate that these differences may be related to differences in pathology between glaucoma and LHON. Specifically, while visual field damage is specific to the fovea in LHON ^76^, glaucoma patients often exhibit damage within a wide range of visual fields. Therefore, such differences in visual field damage may explain the differences in diffusivity along the OT. In addition, the differences in disease progression may affect diffusivity along the OT, because while dendrite and soma damage precede axonal damage in glaucoma ^77–79^, it is unclear whether a similar order of progression occurs in LHON.

This study has several limitations. First, although the sample size used in this study was sufficient to identify large group differences between glaucoma and control subjects, statistical power was still limited when evaluating correlations between MRI measurements and other types of measurements (such as visual field test) in glaucoma patients. Second, since this study focused entirely on open-angle glaucoma, the results may not be fully generalizable to other types of glaucoma populations ^80^. Third, while the NODDI metrics and qT1 are hypothesized to be relatively specific markers of microstructural properties, they do not have a complete one-to-one relationship with the properties of myelin or axons ^25,81^. A possible way to overcome this limitation is to build a model to predict the fractions of certain types of lipids from multiple quantitative MRI metrics ^45^.

Finally, it is important to understand the extent to which MRI-quantifiable white matter tissue changes are reversible. This requires a longitudinal MRI study to evaluate how specific treatment strategies affect MRI-based metrics. While such a study is not trivial to perform, we hope that the extension of a quantitative multi-contrast MRI approach will continue to improve our understanding of white matter consequences of glaucoma.

## Data Availability

The data and code for reproducing figures and statistical analyses will become available in a public repository after an acceptance of this work in a peer-reviewed journal.

## Acknowledgments

We thank Yusuke Sakai for supporting the data analysis.

## Supporting information of

**Supplementary Figure 1.**
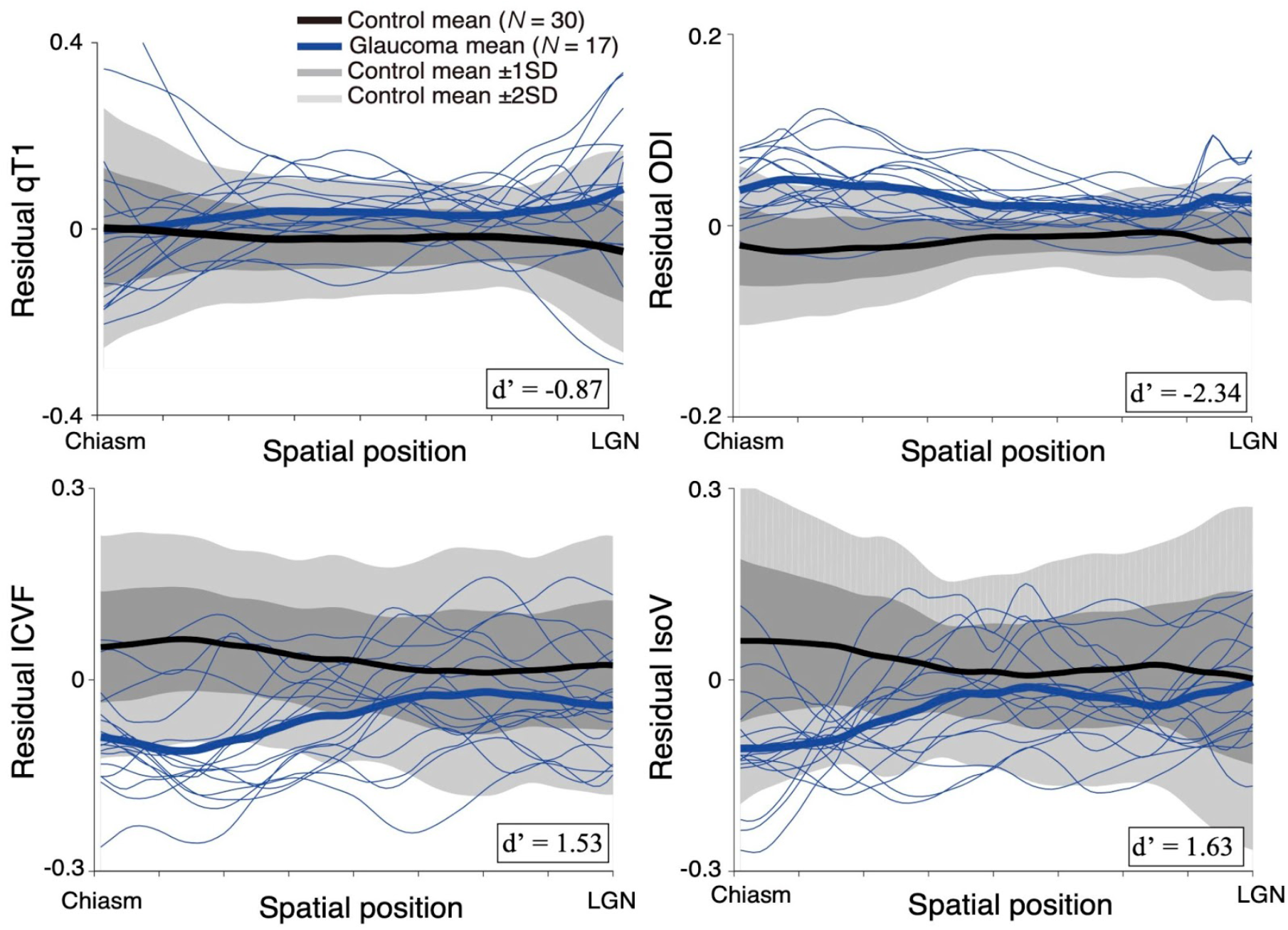
Tissue properties along the OT after using linear regression to control for inter-subject variability in age (see Materials and Methods). The vertical axis depicts the residual of each metric (qT1, ICVF, ODI, and IsoV), which describes the inter-subject variance not explained by age variability. Other conventions are identical to those in Figure 3.

**Supplementary Figure 2.**
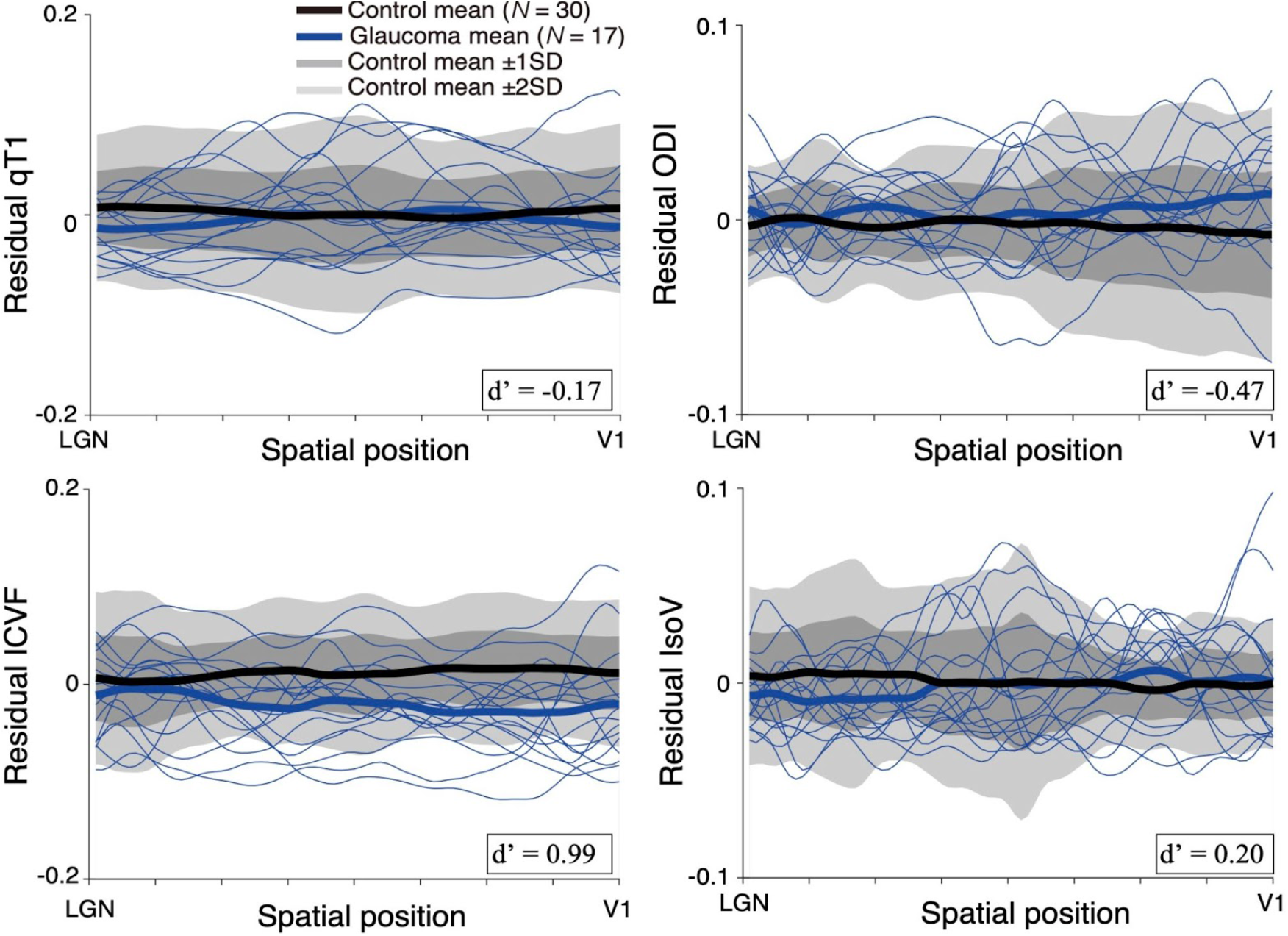
Tissue properties along the OR after using linear regression to control for inter-subject variability in age (see Materials and Methods). Other conventions are identical to those in Supplementary Figure 2.

**Supplementary Figure 3.**
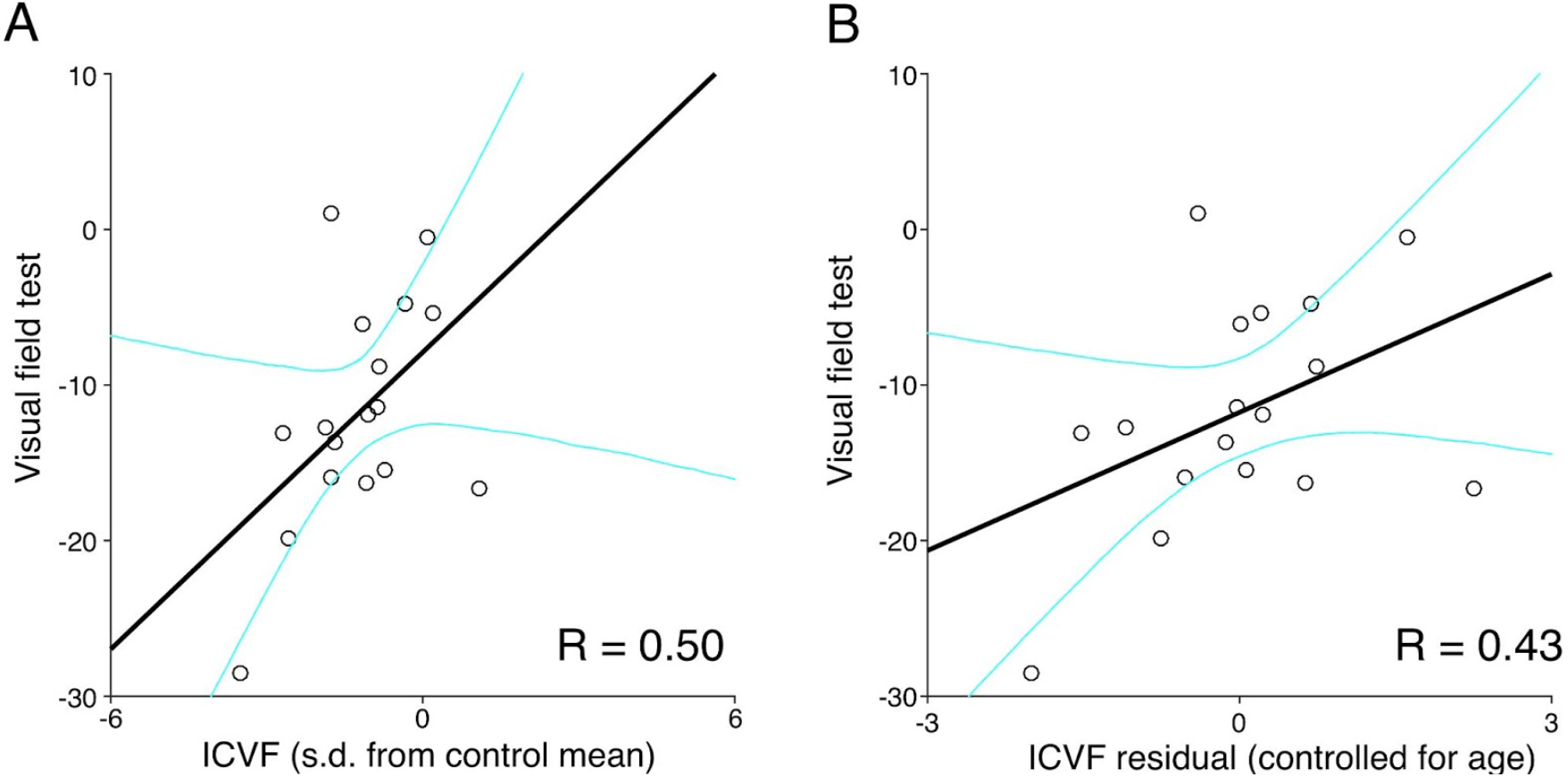
MRI measurements along the OR and results of the visual field test. **A**. Correlation between ICVF along the OR and the visual field test results in glaucoma patients. **B**. Correlation between ICVF along the OR controlled for patient age and the visual field test results.

**Supplementary Figure 4.**
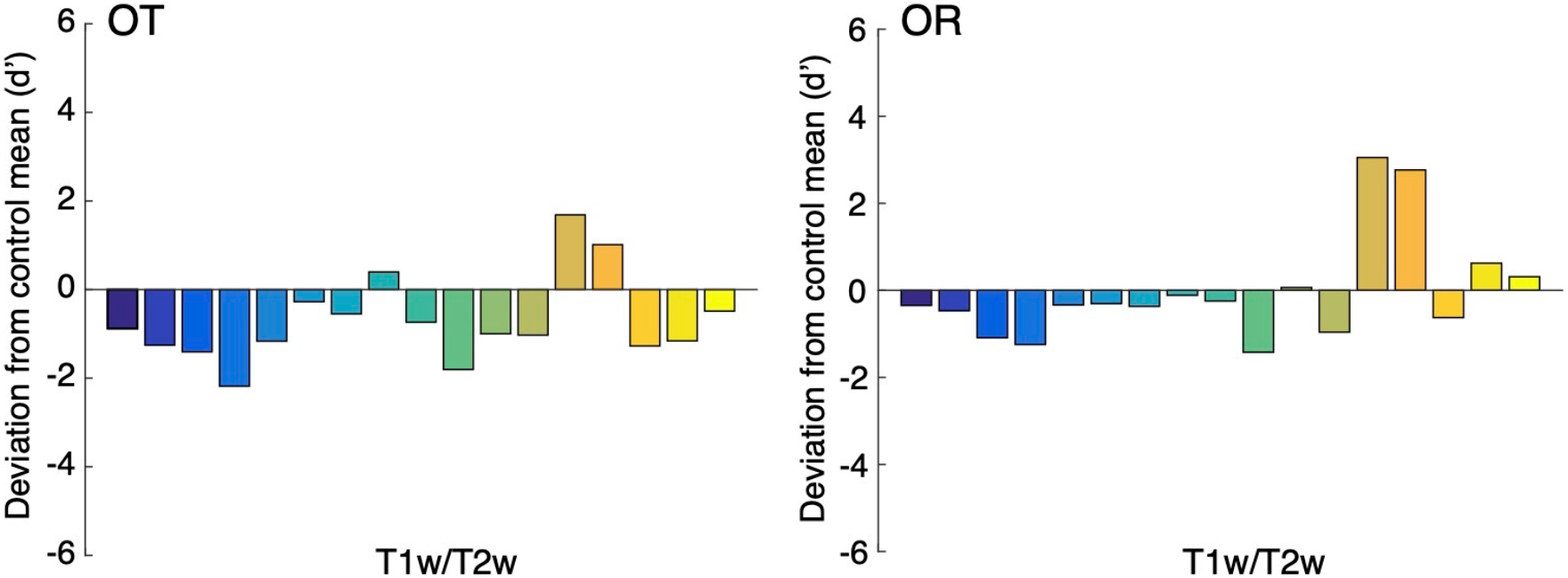
T1w/T2w ratio measurements in glaucoma patients. *Left panel*: OT. *Right panel*: OR. The vertical axis of each plot represents the extent to which individual glaucoma patients (color bars; Glc001-017) deviated from the control mean (*N* = 25). Conventions are identical to those used in Figure 6.

